# Integrated Machine Learning Approaches Highlight the Heterogeneity of Human Myeloid-Derived Suppressor Cells in Acute Sepsis

**DOI:** 10.1101/2022.07.25.22278014

**Authors:** Anthony S. Bonavia, Abigail Samuelsen, E. Scott Halstead

## Abstract

Highly heterogeneous cell populations require multiple flow cytometric markers for appropriate phenotypic characterization. This exponentially increases the complexity of 2D scatter plot analysis and exacerbates human errors due to variations in manual gating of flow data. We describe a workflow involving the stepwise integration of several, newly available machine learning tools for the analysis of myeloid-derived suppressor cells (MDSCs) in septic and non-septic critical illness. Unsupervised clustering of flow cytometric data showed good correlation with, but significantly different numbers of, MDSCs as compared with the cell numbers obtained by manual gating. However, both quantification methods revealed a significant difference between numbers of PMN-MDSC at day 1 in healthy volunteers and critically ill patients having septic or non-septic illness. Numbers of PMN-MDSC obtained by machine learning positively correlated with 30 days hospital readmission following critical illness, whereas manual gating of this cell population distinguished between septic and non-septic critical illness. Neither gating strategy found a correlation between number of MDSCs and 30-day mortality or hospital length of stay.

## Introduction

Myeloid-derived suppressor cells (MDSCs) are a heterogeneous population of myeloid cells that suppress T cell and natural killer cell activity. Previously known as “natural suppressor” cells, these cells are believed to be central to the pathogenesis of cancer, where they cause immune dysfunction and resilience to chemotherapeutic agents (1, 2). More recently, MDSCs have also been implicated in the pathophysiology of sepsis, the life-threatening organ dysfunction resulting from a dysregulated host immune response to infection (3, 4). However, MDSCs may also play a protective role during hyperinflammatory disease processes (5).

The rapid progression of sepsis leaves a narrow but critical time window in which clinicians can potentially intervene to improve patient outcomes. MDSCs have been proposed as a therapeutic target in this time window (3), although investigations into their pathophysiology have met several challenges. First, the gold standard for quantifying MDSC number and/or function involves a T cell proliferation assay which measures their ‘suppressive’ activity over several days. This delay impacts our ability to intervene early in sepsis via the administration of immune adjuvants to patients who may benefit from this therapy. Second, sepsis is a highly heterogeneous syndrome marked predominantly by hyper-inflammation in certain patients, immune paralysis in others, but often by both processes in concert (6, 7). It is likely that both MDSC number and function are equally heterogeneous in these different sepsis subtypes. Murine MDSCs are ubiquitously identified as CD11b^+^ Ly6G^+^ Ly6C^lo^ (PMN-MDSC) or CD11b^+^ Ly6G^-^ Ly6C^hi^ (M-MDSC). In contrast, there is tremendous inconsistency in the nomenclature and surface markers that characterize human MDSCs. Despite general guidelines designed to minimize bias in the presentation and interpretation of flow cytometric data, significant variations between individuals and laboratories persist and may affect the conclusions drawn from flow cytometry data (8, 9).

We hypothesized that unsupervised machine learning algorithms would highlight MDSC subpopulations, on day 1 of critical illness, that correlate with patient outcomes. We investigated our hypothesis by applying a stepwise, unsupervised clustering workflow for quantifying MDSC, and we then compared results obtained with those using manual gating alone. We subsequently examined the relationship between the number of MDSCs calculated by each method to patient outcomes. Our approach was designed with particular attention to minimizing the introduction of human bias in the identification of MDSCs, by leveraging machine learning for most of the processing of flow cytometric data. The significance of this standardized approach is that it could potentially be applied to prospective clinical trials targeting highly heterogeneous cell populations, whether sepsis-related or not.

## Results and Discussion

Our study cohorts consisted of 17 critically ill and septic (CIS) patients, 5 critically ill and non-septic (CINS) patients and 5 healthy volunteers (**Table 1**). The mean age of healthy volunteers was 43 years (range 24-63) and 80% were male. One was Asian, another Hispanic and three Caucasian. Conventional, manual gating of flow cytometric data was first performed (**Fig 1A)**. Machine learning and optimized gating for PMN-MDSC, M-MDSC and eMDSC was run in parallel by using the strategy described in **Fig 1 B-H**. FlowSOM identified a single e-MDSC metacluster, two M-MDSC metaclusters and ten PMN-MDSC metaclusters (**Fig 1D**). When this data was integrated with dimensionally reduced data produced by UMAP (**Fig 1E**), the integrated model identified one additional M-MDSC metacluster and eliminated two of the previously identified PMN-MDSC clusters (**Fig 1F**). Each MDSC cluster was then processed in Hyperfinder (**Fig 1G**), which reported gating F-measures of 0.932 for e-MDSC, 0.931 ± 0.039 for M-MDSC and 0.860 ± 0.065 for PMN-MDSC.

**Table 1:**
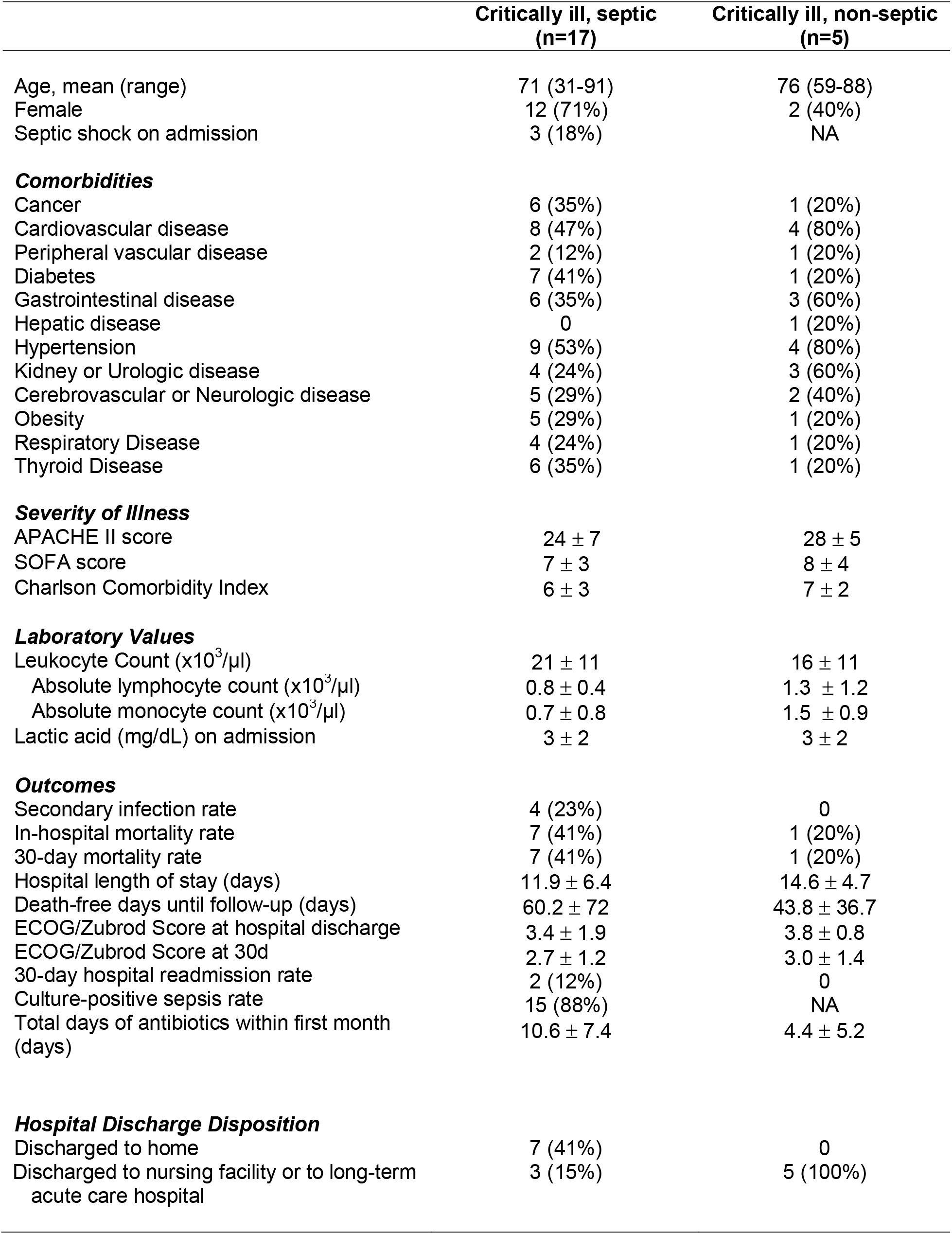
Patient Demographics and Outcomes

**Fig 1.**
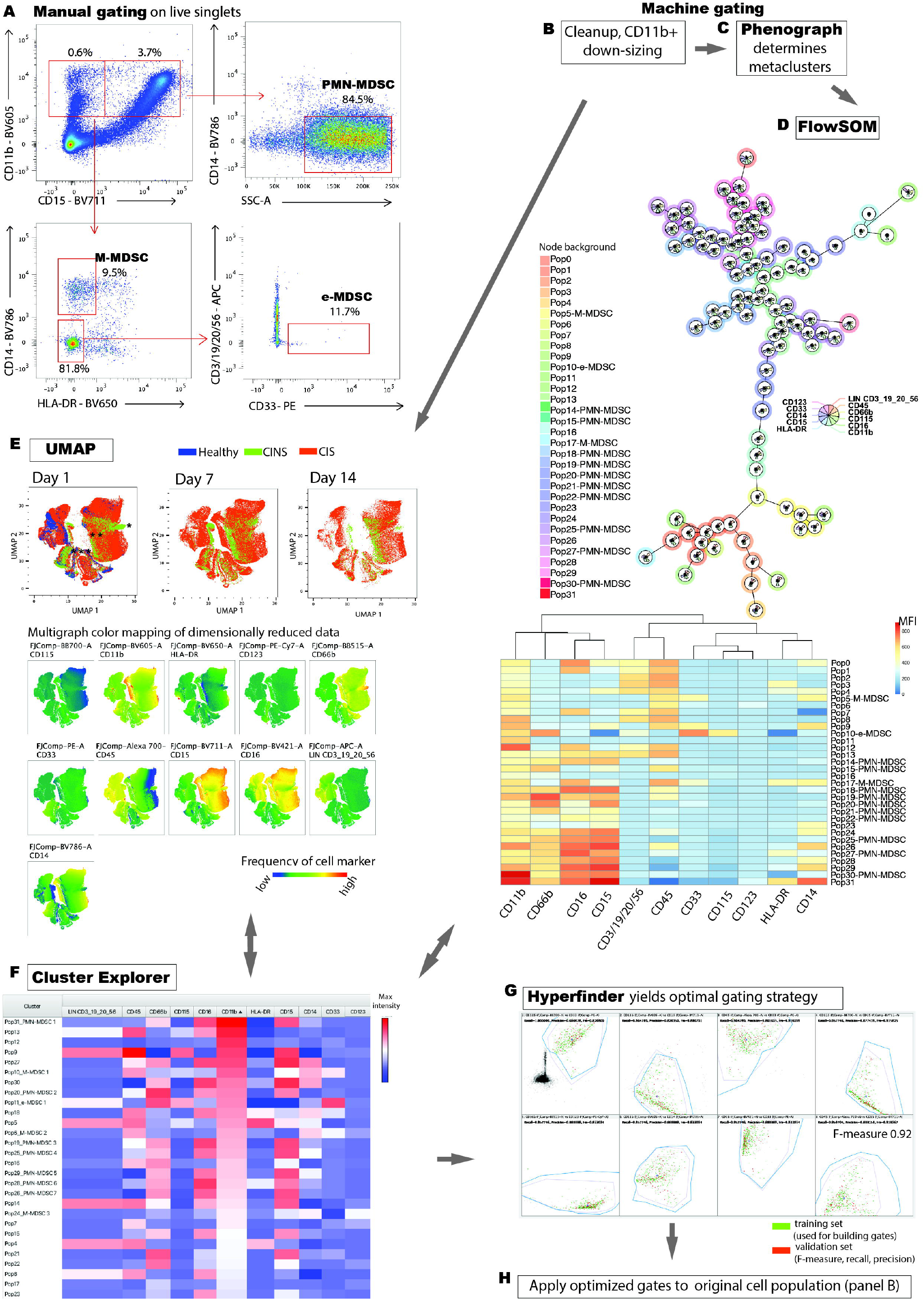
Manual versus machine gating of MDSC populations. **(A)** Representative manual gating strategy used to identify MDSC. M-MDSCs are CD11b+, HLA-DR-, CD15-, CD14+; PMN-MDSCs are CD15+, CD14-, CD11b+, SSC^hi^; e-MDSCs are CD14-, CD15-, CD3-, CD19-, CD20-, CD56-, HLA-DR-, CD33+, CD11b+. **(B)** Stepwise integration of unsupervised, machine learning techniques facilitates the unbiased analysis of high parameter flow cytometry data. Rudimentary gating isolates viable singlets, followed by manual gating for the ‘lowest common denominator’ (CD11b surface marker, in this case) enriches the population of interest. Downsizing to a minimal, common cell count eliminates sampling bias. **(C)** Phenograph identifies the number of metaclusters based on cell markers of interest. **(D)** This number is then inputted into FlowSOM, which generates a minimal spanning tree. PMN-MDSC, M-MDSC and e-MDSC are user-defined annotations based on the definitions in A. **(E)** Dimensionality reduction allows two-dimensional visualization while preserving global data structure. Proximal clusters on the 2-dimensional plot have closely related surface marker profiles. UMAP allows user-directed validation of the clusters identified by FlowSOM and Phenograph. *denotes potential M-MDSC, **denotes potential PMN-MDSCs, ***denotes potential e-MDSCs. **(F)** FlowSOM and UMAP data are integrated in Cluster Explorer, allowing in-depth data exploration. Heatmap illustrates FlowSOM clusters, sorted by increasing intensity of CD11b+ expression. Numeric annotations denote intermediate phenotypes as determined by authors. Note that the integration of data generated using different machine learning tools re-defines certain MDSC clusters, as compared with results from D or E alone. **(G)** Hyperfinder optimizes polygon gating of populations of interest. **(H)** The generated algorithm is then iteratively applied to flow data generated in B. Displayed is an example of Hyperfinder gating for cluster e-MDSC-1.

The numbers of PMN-MDSC as assessed by machine gating correlated with, but were significantly different from, those identified by manual gating (P=0.05, paired t-test), as were numbers of e-MDSC (P<0.0001) and M-MDSC (P=0.003) (**Fig 2A**). This data, in conjunction with a decreased F score for PMN-MDSC, highlights the difficulty in classifying this latter MDSC variant based on surface markers alone. It is probable that a portion of PMN-MDSC thus identified are mature neutrophils, since human CD11b^+^CD15^+^CD14^-^CD33^+/lo^CD66b^+^ markers enrich for neutrophils at all maturation stages (10). Gradient centrifugation using 1.077 g l^-1^ density has been suggested to separate neutrophils from neutrophilic MDSC, since the latter are enriched in low density (mononuclear cell fraction) whereas neutrophils are high density cells. However, cross-contamination of fractions is a common, recognized and unavoidable problem whether density gradient centrifugation is utilized or not (11). Rather than provide precise quantification of PMN-MDSC, our study is intended to illustrate the potential applications of machine learning approaches in multiparameter analysis of phenotypically heterogeneous cell populations.

**Fig 2.**
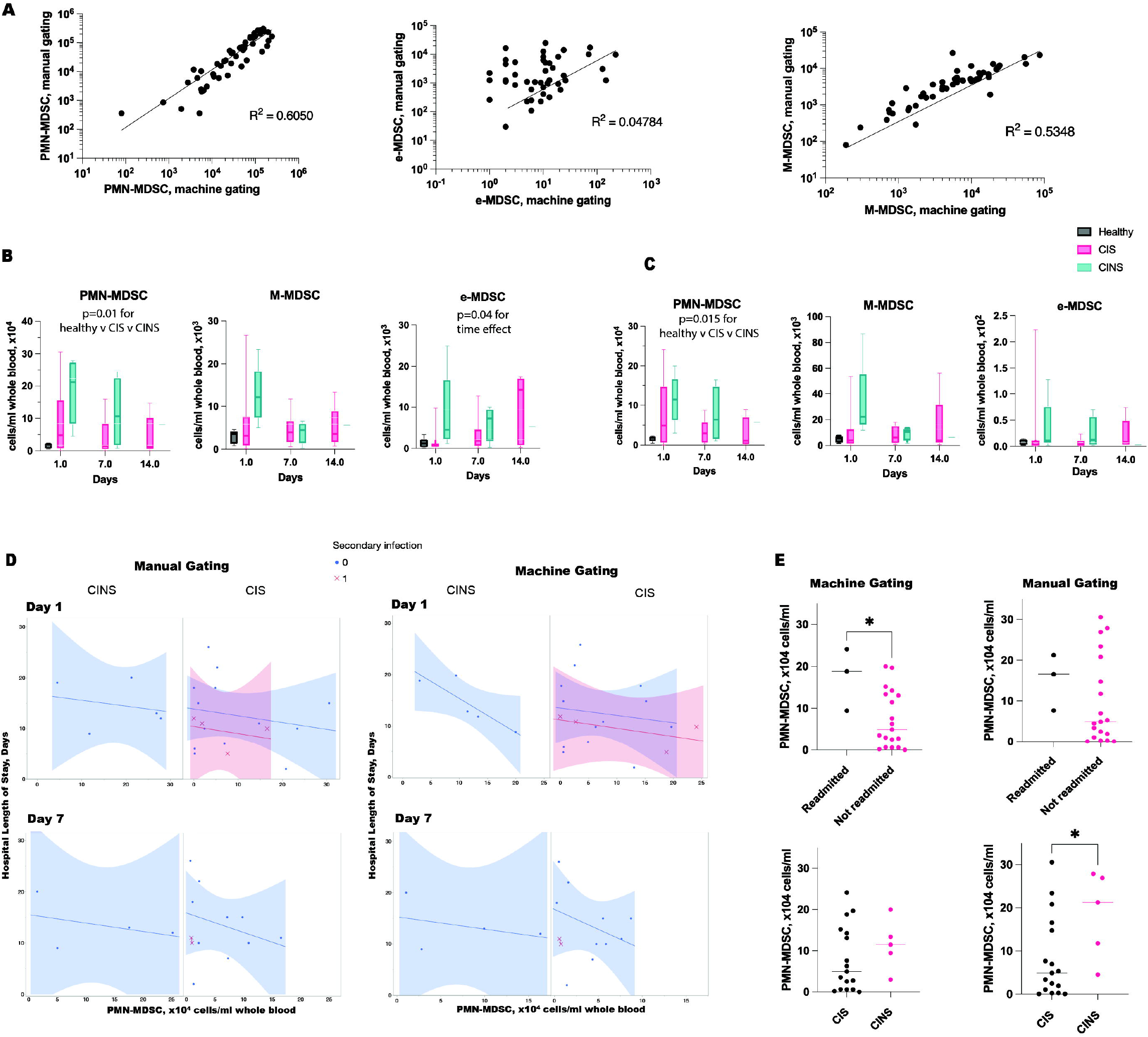
MDSCs are expanded during septic and non-septic critical illness. **(A)** Correlation between MDSC populations by manual gating versus machine learning. Boxplots (Tukey) representing normalized cell counts of PMN-MDSCs, M-MDSCs, and e-MDSCs at different time points in critically ill and septic (CIS, N = 17), critically ill and non-septic (CINS, N = 5) and in healthy donors (N = 5) by manual gating **(B)** and machine learning **(C). (D)** Number of PMN-MDSC at day 1 and day 7 are negatively correlated with hospital length of stay and risk of secondary infections. Line of fit shows linear regression with confidence intervals. **(E)** Increased numbers of PMN-MDSC at day 1, by machine gating, are associated with 30-day hospital readmission (P = 0.03), whereas PMN-MDSC on day 1, by manual quantification, correlate with a diagnosis of sepsis (P = 0.05).

Mixed, main effects analysis of both manual and machine gating of MDSC populations identified a significant difference in total PMN-MDSCs between healthy, CIS and CINS patients on day 1 of study enrollment (P=0.01 for manual gating, P=0.015 for machine learning) (**Fig 2B and 2C**). There was no significant difference between CIS and CINS in the number of e-MDSC or MDSC, or in the total number of PMN-MDSC at 7- and 14-days post-study enrollment. While our CINS group consisted of only five patients, this finding suggests that critical illness and multi-organ dysfunction (rather than sepsis) may be the primary cause of MDSC expansion. Moldawer *et al* have proposed that sepsis-induced chronic critical illness results from proliferation of MDSC (12), and that this cell population drives the persistent inflammation-immunosuppression and catabolism syndrome that commonly follows prolonged surgical sepsis (13). However, it is also possible that stress and inflammation associated with critical illness are drivers of this process, perhaps contributing to the elevated rates of nosocomial infections observed in patients experiencing prolonged stays in critical care wards.

In our cohort, none of the MDSC cell populations (PMN-MDSC, M-MDSC or e-MDSC) evaluated by manual or machine gating predicted 30-day mortality. Neither did any individual sub-population predict mortality, leading us to accept our null hypothesis. MDSC number did not correlate with severity of illness by APACHE II, SOFA scores or Charlson Comorbidity Indices. The number of PMN-MDSC did correlate with hospital length of stay and risk of secondary infection, although no one sub-population of PMN-MDSC alone could explain this difference (**Fig 2D**). Increased numbers of PMN-MDSC at day 1 predict hospital readmission (machine gating, P=0.03) and a diagnosis of sepsis (manual gating, P=0.05) (**Fig 2E**).

The main limitation of our study is that we cannot, with certainty, conclude that the analyzed cell populations constitute ‘true’ MDSCs without ascertaining that they suppress T cell proliferation *in vitro*. Conversely one cannot quantify suppressive potential without first identifying and sorting cells according to their surface markers. The importance of assessing suppressive MDSC cell activity may be further obviated by evidence that, while sepsis survivors have elevated numbers of MDSCs for at least 6 weeks following infection, only MDSCs that are obtained at >14 days post-sepsis suppress T lymphocyte proliferation and IL-2 production (13). The latter study suggests that, unlike murine studies or studies *in vitro*, human MDSCs produced early in sepsis may have not yet adopted immunosuppressive properties, further increasing the importance of accurate flow cytometric quantification of these cell populations for future investigation.

## Concluding Remarks

The integration of machine learning tools into flow cytometric software minimizes both human processing time and intuition-based analysis of data. It also excels in the quantification of cell populations with *intermediate* phenotypes. Sepsis is a syndrome that often has a nebulous onset and rapid clinical evolution, and we demonstrate that the proportions PMN-MDSC correlate with secondary infection and hospital readmission, although no one sub-population that was identified by machine learning can explain these differences. The strategy we describe may be particularly useful when employing multiple flow parameters in the investigation of highly heterogeneous cell populations. It utilizes readily available tools and thus has a low investigator barrier to entry. It may also be applied to the cellular analysis of several, non-sepsis diseases. Its stepwise, algorthimic nature may be beneficial in multi-institutional research endeavors as an alternative to the transfer of research samples to a centralized facility for standardized analysis.

## Methods

### Study Participants

This prospective, observational trial was performed on critically ill, adult patients and healthy control volunteers, between 11/2021 and 6/2022. All participants provided informed consent in accordance with the institutional Human Study Protection Office (#15328,#10357). Sepsis was defined as a change in sequential organ failure assessment (SOFA) score of two or more in the setting of clinically suspected or microbiologically proven infection (4). Critical illness was defined as the need for continuous intravenous infusion of vasopressors to maintain a mean arterial pressure of ≥65 mmHg, and/or the need for continuous respiratory support and monitoring, and/or the need for continuous renal replacement therapy. Critically ill and non-septic patients included adult patients who were older than 18 y and fulfilled criteria for critical illness but not sepsis. Healthy volunteers included adults who did not fulfil criteria for critical illness or sepsis. To minimize the potential or confounding effects, we excluded patients with active hematologic malignancies, autoimmune disorders and those who were receiving immunomodulating therapies.

### Clinical variables

Patient data for critically ill, septic (CIS) and non-septic (CINS) patients was obtained from the electronic medical record. To distinguish between illness severity in patients, we utilized the Charlson Comorbidity Index (14), the Acute Physiology and Chronic Health Evaluation (APACHE II) and Sequential Organ Failure Assessment (SOFA) scores (15–17).

### Processing of blood samples

Venous blood samples were collected in tubes containing ethylenediamine tetra-acetic acid (EDTA), within 24h of the onset of critical illness +/- sepsis (day 1) and on subsequent days 7-10 and 14, if the patient was still alive and hospitalized at that time. 100µl of whole blood was blocked with mouse serum (0.5:1, M5905, Sigma-Aldrich, St. Louis, MO), for 5 minutes at room temperature followed by addition of e780 Fixable Viability Dye (1:800, cat #65-0865, eBioscience, San Diego, CA). The following antibodies were then added for 20 minutes at room temperature, in the dark: anti-CD66b-FITC (1:20, cat #555724, BD Biosciences, San Diego, CA), anti-CD115-PerCP-Cy5.5 (1:200, clone 9-4D2-1E4, #347309, Biolegend, San Diego, CA), anti-CD16-BV421 (1:80, clone 368, #562874, BD), anti-CD11b-BV605 (1:80, clone ICRF44, #301332, Biolegend), anti-HLA-DR-BV650 (1:80, clone L243, #307649, Biolegend), anti-CD15-BV711 (1:80, clone W6D3, #563142, BD), anti-CD14-BV786 (1:80, clone M5E2, #563699, BD), anti-Lineage Cocktail CD3/19/20/56-APC (1:20, #363601, Biolegend), anti-CD45-AF700 (1:80, clone 2D1, #368513, Biolegend), anti-CD33-PE (1:20, #555450, BD), anti-CD123-PE-Cy7 (1:80, clone 7G3, #560826, BD). This antibody panel was created following a literature search into recently published strategies for identifying human MDSCs (12, 13, 18–20).

Following antibody staining, red blood cells were lysed and cells were washed and fixed. Counting Beads (Invitrogen, Waltham, MA) were added to a separate sample of lysed, unstained cells. Analysis was performed on FACSymphony A3 (Becton Dickinson & Company, Franklin Lakes, NJ) and using Flowjo v10.8.1 (BD Biosciences).

### Manual gating of flow cytometric data

MDSC nomenclature was consistent with recently described minimal phenotypic characteristics necessary to identify cells as MDSC (11) and with nomenclature utilized in a comparable study of septic patients (20). M-MDSCs were CD11b^+^, CD15^−^, CD14^+^, HLA-DR^−^; PMN-MDSCs were CD15^+^, CD14^−^, CD11b^+^, SSC^hi^; e-MDSCs were CD3^−^, CD14^−^, CD15^−^, CD19^−^, CD56^−^, HLA-DR^−^, CD33^+^, CD11b^+^. Flow data was analyzed and manual gating performed and presented in accordance with guidelines for the use of flow cytometry in immunologic studies (8) (**Fig 1A**).

### Machine gating followed by unsupervised clustering of flow cytometric data

A comprehensive literature search revealed that the only between-study consistency in human MDSC characterization is the pan-myeloid, CD11b^+^ surface marker. Thus, we pre-processed flow data by applying bead compensation and then manually gating for viable, CD11b^+^ singlets by using Flowjo 10.8.1 (BD Biosciences) (**Fig 1B**). We employed a ‘lowest common denominator’ approach to minimize bias, to preserve integrity of the dataset and to rely predominantly on machine learning for subsequent exploration. 2D scatter plots clearly delineated between CD11b^-^ and CD11b^+^ populations in all samples. We then used the Flowjo Downsampler tool to export an equal number of normally distributed CD11b^+^ cells from each sample for subsequent processing.

Unsupervised clustering was performed by two independent methods. PhenoGraph discovers subpopulations by using nearest-neighbor graphing, wherein each cell is represented by a node (in high-dimensional space), connected by a set of edges, to a neighborhood of its most similar cells (21) (**Fig 1C**). The graph distils the high-dimensional distribution of single cells into a compact, information-rich data structure that captures phenotypic relatedness and overcomes many pitfalls of standard geometries (21). The strength of PhenoGraph is its rapidity and the unsupervised determination of cluster number, although its weakness is the lack of visual output that illustrates the relatedness of subpopulations. The number of Phenograph-identified metaclusters was inputted into FlowSOM, a self-organizing map algorithm in which each node represents a point in the multidimensional input space, and each new point is classified with the node that is its nearest neighbor (22) (**Fig 1D**). The grid contains topological information about the relatedness of each point, such that nodes closely connected to each other on the grid resemble each other more closely than those connected through a long path. FlowSOM provides visual representation of data by using a minimal spanning tree, although it requires the user to pre-define a maximum number of metaclusters, leading to potential introduction of user bias. Hence the use of Phenograph to identify cluster number, prior to FlowSOM.

Uniform Manifold Approximation and Projection (UMAP) was used for dimension reduction of data and alternate clustering (23) (**Fig 1E**). UMAP is a recently introduced, scalable machine learning algorithm that creates an intuitive, 2-dimensional map wherein spatial proximity of clusters implies similar cell marker phenotype.

### Comparison of Clusters, Automated Gating and Iterative Application to Samples

ClusterExplorer allows the selection and integration of one or more clustering parameters (in this case, FlowSOM and Phenograph-generated clusters) with user-defined surface markers of interest and dimensionality reduction x and y parameters (in this case, by UMAP), on which clusters can be displayed. We selected ClusterExplorer heat map as the primary tool with which identify discrete MDSC populations, based on the intensity of different surface markers as they relate to characterization of each of the three MDSC types (**Fig 1F**). However, ClusterExplorer can presents data on profile graphs with relative expression levels of surface markers and barcharts showing the relative numbers of events in each cluster. Notably, this step was the only one, in the machine gating algorithm, that involveed investigator input, and therefore the machine learning algorithm was not completely free from potential investigator error.

After identifying MDSC subsets by using ClusterExplorer, Hyperfinder was used to optimize the gating strategy for each identified cell population (maximum of 8 gates, target F-measure beta of 1) (**Fig 1G**). F-measure is the harmonic mean of yield and purity, with a score of 1 indicating equal contributions of yield and purity. The resulting gating algorithm for each MDSC subset was then retroactively applied to the original CD11b^+^ population from each sample (**Fig 1H**). The number of cells identified by this approach that met criteria for eMDSC, M-MDSC and PMN-MDSC was then compared with equivalent populations of cells identified by manual gating.

### Statistical Analyses

Analyses were performed in Prism v.9.3.1 (Graphpad Software, San Diego, CA) and JMP Pro 16.0.0 (SAS Institute Inc., Cary, NC). Details of each analysis are contained within the table and figure legends.

## Data Availability

All data produced in the present study are available upon reasonable request to the authors

## Abbreviations

APACHE: Acute Physiology and Chronic Health Evaluation
e-MDSC: early myeloid-derived suppressor cells
MDSC: myeloid-derived suppressor cells
M-MDSC: monocytic myeloid-derived suppressor cells
PMN-MDSC: neutrophilic myeloid-derived suppressor cells
SOFA: Sequential Organ Failure Assessment
UMAP: Uniform Manifold Approximation and Projection

## Author Contributions

ASB: Conceptualization, Methodology, Formal Analysis, Funding Acquisition, Investigation, Writing Original Draft, Review and Editing, Project Administration. AS: Investigation, Data Curation, Writing - Review and Editing. ESH: Formal Analysis, Writing Original Draft, Reviewing and Editing.

## Data availability

All data are available by direct communication with the authors.

## Conflicts of Interest

The research was conducted in the absence of any financial relationships that could be construed as a potential conflict of interest.

## Acknowledgements

Funding was provided by the National Institute of General Medical Sciences #K08GM138825 (ASB).

## Notes

### Competing Interest Statement

The authors have declared no competing interest.

### Funding Statement

Funding was provided by the National Institute of General Medical Sciences, grant #K08GM138825 (ASB).

### Author Declarations

All patients were enrolled at the Penn State Milton S Hershey Medical Center (Hershey, PA), following study approval by the Human Study Protection Office (Institutional Review Board Approval #15328 and #10357). Informed consent was obtained from patients having decision-making capacity, or the legally authorized healthcare representatives of patients lacking decision-making capacity.

